# Hand sanitizer recalls during the COVID-19 pandemic: a descriptive analysis of regulatory records from Canada, Nepal, Singapore and the USA

**DOI:** 10.64898/2026.10.26.26366567

**Authors:** Ramdeep Shrestha, Kriti Khanal, Astha Neupane, Basant Giri

## Abstract

**Background:** Alcohol-based hand sanitizers were central to infection prevention during the COVID-19 pandemic. The abrupt rise in global demand fueled the emergence of substandard and unregulated products. Government regulatory bodies around the globe responded to this issue by tightening quality control and enforcing stringent limits on contaminants. This study aimed to descriptively analyze the reasons for, and timing of hand sanitizer recalls and safety warnings between 2020 and 2024.

**Methodology:** We extracted publicly available records from the national regulators of Canada, Nepal, Singapore, and the USA. The database was analyzed by country, year, and reason for recall to identify the recall patterns and regulatory practices.

**Results:** We identified 1044 recalled products in Canada (277), Nepal (39), Singapore (26), and the USA (702). Major reasons for recall were administrative reasons such as non-compliance, non-response to the regulator, and unregistered or unauthorized sale (43.9%) followed by contamination with hazardous chemicals such as methanol, acetaldehyde, and benzene (37.4%) and subpotent ethanol or isopropanol (15.6%). Two-thirds of the recall records were from the USA followed by Canada, Nepal, and Singapore. While most of the recalls were announced during the first year of COVID-19, the recalls did not follow a temporal trend.

**Conclusion:** The recall records showcased product hazard risks during the pandemic, highlighting its global impacts. The recalls in each of the four countries may have missed other substandard hand sanitizer products. This underlines the importance of strict monitoring and public awareness for product safety. The recall records discussed in this paper describe regulatory activity and should not be read as the prevalence of substandard hand sanitizers. Robust emergency regulatory frameworks, rapid analytical verification, and distributed yet strictly monitored certified production systems may help reduce public health risks due to hand sanitizers in the future.

## Introduction

Hand sanitizers offer a rapid and practical solution for the prevention of infection spread in healthcare and community settings.^1^ Generally, there are two broad groups of hand sanitizers: alcohol-based hand sanitizers (ABHS) and non-alcohol-based hand sanitizers (NABHS).^2,3^ During the COVID-19 pandemic, the World Health Organization (WHO), the Centers for Disease Control and Prevention (CDC), and the US Food and Drug Administration (FDA) consistently recommended the use of ABHS containing at least 60% ethanol or 70% isopropanol because of their effectiveness in deactivating SARS-CoV-2 and many other pathogens.^2–4^ ABHSs are less effective against some non-enveloped viruses such as norovirus, and hands that are visibly soiled should be washed with soap and water.^5^ Alcohol denatures proteins and disrupts microbial membranes, and the water fraction (10-30%) slows evaporation, which maintains the contact time needed for activity.^6^ NABHSs, for example those based on benzalkonium chloride, act as cationic surfactants and leave a residual antimicrobial film. They are gentler on skin but are generally less effective than ABHSs. ^5,7^

The demand for hand sanitizers surged unprecedentedly following the WHO’s declaration of COVID-19 as a worldwide pandemic on 11 March 2020^8^. Market analysis estimates that global hand sanitizer revenue rose from USD 1.03 billion in 2019 to USD 6.3 billion in 2020 and has since settled near USD 3 billion.^9^ The market is projected to reach USD 4.7 billion by 2034. North America only held 40.45% of the market in 2025. Canada reported a rise in sanitizer sales of more than 600% between 2019 and 2020.^10^ The hand sanitizer growth was over 250% in the U.K., 1800% in Italy, and monthly growth over 1,380% in the U.S. from December 2019 to January 2020. Nepal also saw rapid growth in demand, attributed to awareness regarding the usefulness of the sanitizer.^11^

The global high demand was met partly by new, inexperienced, or unregulated manufacturers, leading to substandard, fake, or counterfeit products. Studies reported a significant presence of methanol, benzene, and 1-propanol adulteration in some products.^12,13^ Methanol is of particular concern because ingestion, or to a lesser extent prolonged skin exposure, can cause blindness, neurological damage, and death.^14,15^ Fatal cases of methanol toxicity were reported from the southwest USA^16^. In response, regulators issued several product recalls and warnings and implemented tighter surveillance. They also intensified their efforts to monitor and enforce quality standards to ensure product safety by setting stringent criteria to guarantee the safety of hand sanitizers.^17,18^ For instance, Health Canada’s limit for methanol has been set to 200 ppm while the US-FDA has set it to 630 ppm in sanitizers. ^19,20^ To ensure product safety, contaminants like methanol, benzene, and acetaldehyde are mostly assessed due to their toxicity.

Products that fail to meet safety standards are subject to be recalled through a systematic process^21^. A manufacturer found to be non-compliant is first notified and given the chance to respond. If corrective action is inadequate, a public recall or warning is issued, products are withdrawn from sale, and follow-up testing precedes any return to market. The recall records are readily available, though imperfect, indicators of product quality failures and of regulatory response.

We analyzed official hand sanitizer recall records from Canada, Nepal, Singapore and the USA for 2020 to 2024 to describe (i) recall counts, (ii) the reasons for recall, (iii) differences in reason profiles between countries, (iv) trends over time. We aimed to identify lessons for regulatory preparedness in future public health emergencies.

## Methods

### Study design and data sources

We conducted a retrospective descriptive study of publicly available regulatory records. Records were extracted from four national agencies: Department of Drug Administration (DDA) of Nepal^22^, Health Canada, US FDA, and the Health Sciences Authority (HAS) of Singapore during 2020 to 2024. These countries were chosen to represent two large manufacturing markets with established regulators (USA, Canada), a high-income, import-reliant trading hub with a strict regulatory culture (Singapore), and a low-income, import-dependent market (Nepal). These agencies publish different kinds of records such as recalls, consumer warnings and ‘do-not-use’ listings. These are referred to collectively as ‘recall records’ in this paper.

The time frame of 2020-2024 was chosen to study hand sanitizer recalls because this time frame was characterized by panic buying, emergency manufacturing, and short-term policy overrides leading to multiple quality inconsistencies driven by the COVID-19 global pandemic’s impacts. Subsequently, sanitizer consumption expanded, and intense production was also triggered. Regionally, the USA and Canada represent countries with the largest manufacturing base and strong public health regulations. Singapore, on the other hand, is a quality-sensitive and high-tech market hub which also sped up the homeland production of hand sanitizers alongside imports. ^23^ Likewise, Nepal is an import-dependent and low-income country reflecting the effects in smaller markets.

### Data extraction and classification

For every record, we extracted the product name, the stated reason for recall, the contaminant named (if any), and the date of the notice. Each record was then assigned to one of six categories: (1) toxic chemical contaminants like methanol, acetaldehyde, benzene, or ethyl acetate above the acceptable level; (2) subpotent active ingredient such as ethanol or isopropanol; (3) mislabeling; (4) microbial contamination; (5) administrative reasons such as non-compliance, no response to the regulator, or unregistered and unauthorized sale; and other.

### Data analysis

A thorough and comparative analysis was conducted to identify common recall patterns, key contaminants, and differences in regulatory actions and strategies taken by these countries. We summarized counts by reason, country, and calendar year.

### Ethics

The study used only publicly available regulatory data and involved no human participants.

## Results

### Overview of recall records

A summary of country-wise recall counts and their reasons is provided in Table 1. A complete list of recalled hand sanitizers is given in Table SI 1. We identified a total of 1,044 recall records from Canada (26.5%), Nepal (3.7%), Singapore (2.5%), and the USA (67.2%). Nepal’s DDA issued its first recall notice on April 1, 2020. The US FDA issued a consumer warning about methanol-containing sanitizers on 29 June 2020 and subsequently published a growing ‘do-not-use’ list, which reached 665 products by July 2024. ^24–26^ Health Canada’s first list, published from 17 June 2020 to 24 March 2021, contained 153 products, and a second list during March 2021 – September 2022 added 59 products. ^27,28^ It has not issued any hand sanitizer recall after April 2021 during the COVID-19 time.

**Table 1.** Summary of country wise recall data and reasons for recall.

| Reason for recall | USA | Canada | Nepal | Singapore | Total (% of all) |
| --- | --- | --- | --- | --- | --- |
| **Toxic chemical contaminants** | **254** | **100** | **11** | **26** | **391 (37.5)** |
| Methanol | 202 | 27 | 11 | 8 | 248 (23.8) |
| Acetaldehyde | 30 | - | - | 18 | 48 (4.6) |
| Benzene | 22 | - | - | - | 22 (2.1) |
| Ethyl acetate above acceptable level | - | 73 | - | - | 73 (7.0) |
| **Subpotent ethanol/isopropanol** | **70** | **74** | **19** | **-** | **163 (15.6)** |
| **Mislabeling** | **13** | **7** | **-** | **-** | **20 (1.9)** |
| **Microbial contamination** | **5** | **-** | **-** | **-** | **5 (0.5)** |
| **Administrative: non-compliance, non-response and unregistered or unauthorized sale** | **360** | **89** | **9** | **-** | **458 (43.9)** |
| **Other** | **-** | **7** | **-** | **-** | **7 (0.7)** |
| **Total** | **702** | **277** | **39** | **26** | **1044 (100)** |

*Percentages in the last column are of the 1,044 records. Subtotals for toxic chemical contaminants are shown in the shaded row, with their components indented. – = none recorded.*

### Reasons for recall

Administrative reasons were the most common category overall (458 records, 43.9%) as shown in Table 1. The administrative reason was followed by toxic chemical contaminants (391, 37.5%) and subpotent ethanol or isopropanol (163, 15.6%). Methanol was the most frequent single contaminant (248 records, 23.8%) and was the only contaminant found in all four countries. Other contaminants were acetaldehyde (48 records), ethyl acetate (73, Canada only), and benzene (22, USA only). Mislabeling (20 records) included sanitizers sold in containers resembling water bottles, which raises the risk of accidental ingestion, products with ethanol or isopropanol contents below those claimed, and unsupported claims of effectiveness against COVID-19. Microbial contamination was recorded only in the USA (5 records).

Reason profile differed markedly between countries (Figure 1). In the USA, 51.3% of the 702 records were administrative, almost all because the manufacturer did not respond to FDA notices (360 records). Toxic chemicals accounted for 36.2%, mainly methanol (202 records, 28.8% of US records), acetaldehyde (30), and benzene 922), and subpotency for 10.0% (70 records). In Canada, toxic chemical contaminants (36.1%, methanol 27, ethyl acetate 73), subpotency (26.7%, 74 records), and unauthorized sale 932.1%, 89 records) were of similar importance. The remaining Canadian records involved mislabeling (7) and other reasons (7). In Nepal, subpotency was the leading reason (48.7%, 19 records), followed by methanol (11; 28.2%) and administrative reasons (9; 23.1%). All 26 Singaporean records concerned chemical contaminants – acetaldehyde (18) and methanol (8).

**Figure 1.**
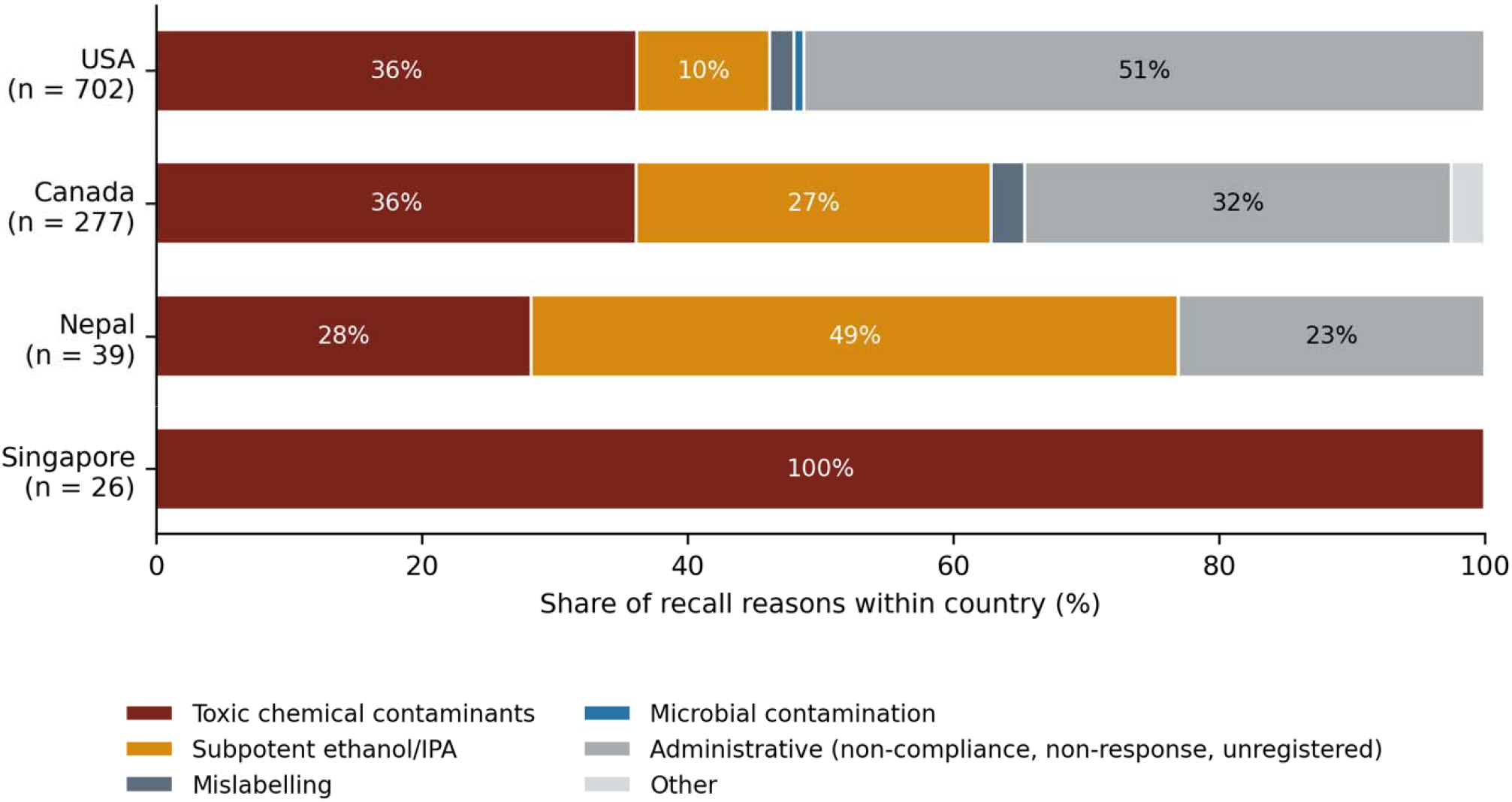
Comparison of recall reasons within each country from 2020 to 2024.

Methanol is a toxic chemical which was found to be improperly substituted for ethanol, especially during peak demand. Such substitutions likely occurred due to shortages of technical-grade ethanol and the emergence of new manufacturers. ^29^ Acetaldehyde is another chemical contaminant which was noted in 30 products in the USA and 18 in Singapore. Similarly, subpotent products due to inadequate ethanol content were also recalled significantly, with 70 cases in the USA, 67 in Canada, and 18 in Nepal. These products failed to meet the minimum ethanol concentration (60-70% v/v) required for effective inactivation of microbes. This indicates a serious inconsistency in product formulation.

Mislabeling was identified in 13 U.S. products and 7 in Canada. Some products were sold in bottles that resembled water bottles, increasing the chances of accidental ingestion. Some claimed to have enough ingredients like ethanol or isopropanol but were found to be subpotent, whereas some products misled consumers with false claims such as “effective against COVID-19”.^30^ Contaminants like Benzene (22 instances in the USA) and ethyl acetate (73 instances in Canada) also indicate the presence of inappropriate solvents.

## Trends over time

The initial year of the pandemic witnessed the highest number of hand sanitizer recalls across all four nations. This peak corresponds with the onset of the pandemic, which spiked demand and resulted in emergency approvals and loosened regulations. During 2021–2022, a general decline in recall incidents was found, which can be attributed to increasing regulatory stabilization, retraction of emergency relaxations, and increased market maturity. Interestingly, the USA experienced a dramatic resurgence in recalls (258) in 2023. It was observed that a huge majority of them (251) recalls were due to the failure of the products to respond to the FDA warnings issued before against various regulatory violations. The rest of the recalls were made for the products produced at the facility that produced acetal- and methanol-contaminated products. This pattern reflects that the regulatory authorities actively monitored throughout the pandemic period. The recalls were mainly due to lapses in industrial compliance and communication rather than regulatory fatigue or emergence of newer counterfeit products. By 2024, recall notices were minimal across all the regions, indicating systemic improvements in production practices and regulatory controls.

The exact number of annual recall notices throughout the years is dispersed, but the recalls persisted with somewhat lower intensity till 2024. Recalls were first recorded in April 2020. The annual pattern differed among the countries (Figure 2). Canada and Nepal recorded their highest annual counts in 2020 (167 and 19, respectively), whereas Singapore’s peak came in 2021 (18 records, after 4 in 2020). In the USA, the annual count fell from 224 in 2020 to 43 in 2021, rose to 107 in 2022, reached its highest value of 258 in 2023, and dropped to 28 in 2024. Thus, the US series does not show a steady decline after the first year.

**Figure 2.**
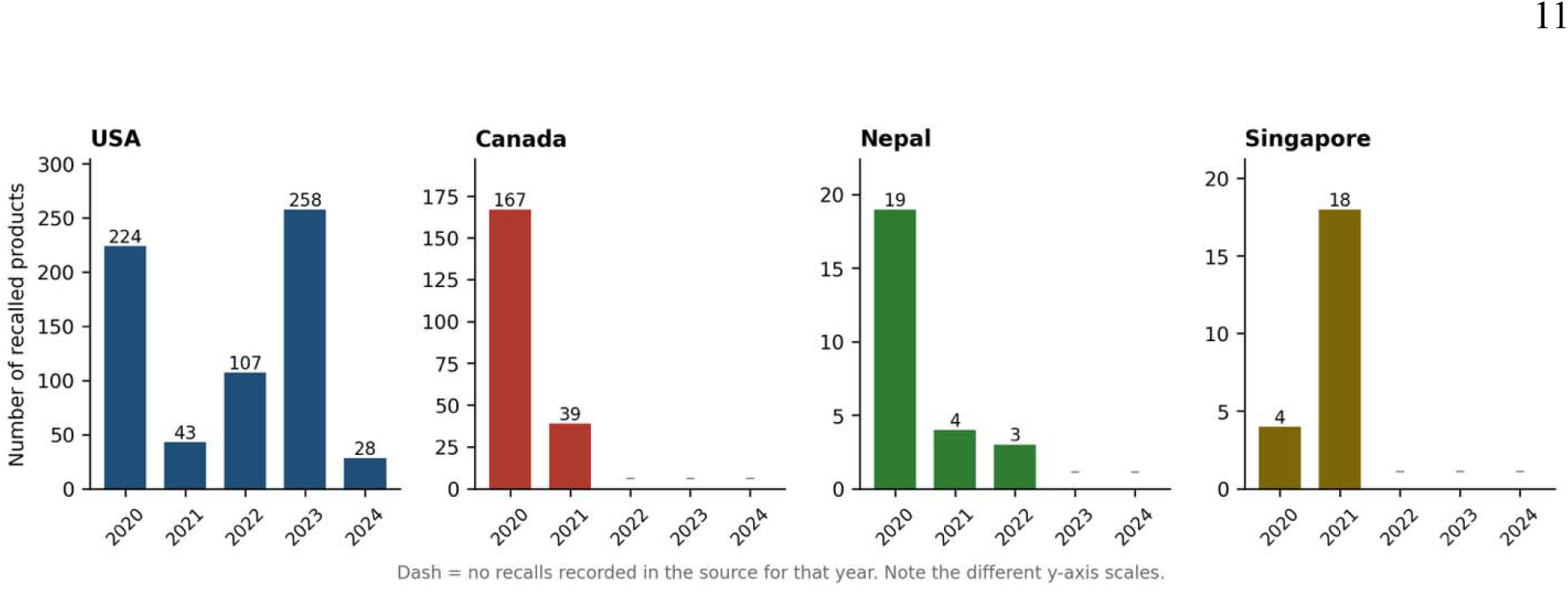
Recall records across four nations from 2020 to 2024.

The 2023 US peak was almost entirely administrative: 251 of the 258 records (97.3%) were for failure to respond to earlier FDA notices, and these 251 records represent 69.7% of all US non-response records. The remaining 2023 records concerned products from a facility that had also made acetaldehyde- and methanol-contaminated products. No Canadian records were dated after 2021, no Nepali records after 2022, and no Singaporean records after 2021. Only the USA has records in 2023–2024.

### Country-wise recalls

The instances of recalls in the USA could be understood as an exemplary case of regulatory overload and non-compliance. The USA reported the highest number of recalls (665) among other countries, of which 360 (over 50% of the total cases) didn’t respond to the FDA notices issued against various reasons. This shows a systemic issue where new manufacturers lack regulatory experience and hence fail to maintain post-market communication. Methanol was the most prominent chemical hazard (202 cases). This incidence also reflects a remarkable regulatory enforcement. Additionally, the notable presence of toxic chemicals like acetaldehyde (30) and benzene (22) poses a serious health hazard upon dermal exposure or inhalation. 70 formulations with subpotent ethanol were also concerning considering the reliance to kill the microbes effectively. This could be alarming, especially in situations like the COVID-19 pandemic where failure of hand sanitizers to kill the virus could result in spread of disease and threat to the lives of many. Besides that, there have been several reports of recalls due to mislabeling and microbial contamination in the marketed products.

In contrast, Canada reveals broader recall reasons with a significant number of cases, reflecting a more rigorous inspection by the authorities. Canada recorded 317 recalls, with significant instances of unauthorized sales (89). The presence of a remarkable number of subpotent ethanol (67), unacceptable levels of ethyl acetate (73) and 1-propanol (7), and counterfeit products (2) underscores both formulation inconsistencies and vulnerabilities in the supply chain. Unlike the USA, Canada’s challenges appear more aligned with formulation and distribution legality rather than communication breakdowns. However, active regulatory actions were observed despite the prevalent systemic production and distribution errors, as marked by non-compliance with the set standards regarding both quality oversight and legality.

Though the number of recalls in Nepal and Singapore was reported on a limited scale; (Nepal:26; Singapore:22), the reasons for recalls were seen to be like those in the USA and Canada. In Nepal, cases of subpotent ethanol (18) and methanol contamination (11) highlight the limited analytical infrastructure available to local manufacturers. The absence of other contaminants was also most likely due to limitations in testing for more complex contaminants and other microbial agents. However, in Singapore, despite its stringent regulatory environment, the presence of acetaldehyde (18) and methanol contamination (8) shows challenges even in tightly controlled markets when demand peaks abruptly. No further hand sanitizer recalls have been issued in Singapore after April 2021, and as such, only data up to 2021 is available for analysis.

Although this study focuses on recall data from the USA, Canada, Singapore, and Nepal, the scope extends beyond domestically manufactured products because of the interconnected global supply chains. The USA, for instance, exports to hundreds of different countries, including China, Canada, Mexico, and Germany, as well as imports from major producers around the globe. Similarly, Canada also trades with many countries. In the case of the Singaporean market, China holds substantial influence, while it also imports hand sanitizers from the European Union, India, Germany, the US, and other nations. ^23^ Due to geographic proximity, a considerable portion of the Nepalese market is occupied by Indian and Chinese products. Therefore, this study also reflects the general trend of hand sanitizer recalls and their reason worldwide.

## Discussion

Since the announcement of the COVID-19 pandemic by WHO in March 2020^31^, several government authorities recommended the use of hand sanitizers. The data suggest that an overwhelming number of recalls were observed during the COVID outbreak, while in the coming years the problem subsided and the recall notices became scarce. Because of quality concerns, the regulatory agencies also started to warn the public and recall certain products. In this analysis of 1,044 hand sanitizer recall records from four countries, two broad problems emerged. The first was product quality. Methanol was found in recalled products in every country, acetaldehyde and benzene in some, and nearly one in six records involved products containing too little alcohol to be effective. The second was regulatory and administrative. The largest single category of records, particularly in the USA and Canada, concerned manufacturers that failed to respond to regulators or sold products without authorisation. The mix of reasons varied significantly between countries, and the timing of recalls differed in each system. Many of the products were recalled because of the violation of different standards at once. For e.g., ‘Subpotent ethanol and presence of methanol’, ‘presence of methanol and acetaldehyde’, or ‘presence of methanol and no response to the authority’.

Methanol is the contaminant of greatest public health concern in our data. The poisonings in Arizona and New Mexico in mid-2020^32^ associated with ingesting hand sanitizer, which caused deaths and permanent visual impairment, show that these recalls addressed real harm. Reports from other countries have likewise described methanol poisoning linked to the pandemic. A plausible explanation is that sanitizer production expanded faster than the supply of pharmaceutical-grade ethanol, so that technical-grade ethanol or ethanol of uncertain origin was used by manufacturers with little experience of quality control^33^. We could not test this explanation, because the records do not report ethanol sources. Acetaldehyde and benzene were less frequent but are also toxic^33^. Acetaldehyde is classified as a probable human carcinogen. Its presence in the ethanol may arise during fermentation, oxidation, or poor purification of ethanol. ^34,35^ The contamination of sanitizers with such aldehydes points out the critical compromise in safety standards during the emergency phase. Benzene is a human carcinogen. Dermal and inhalation exposure from sanitizers has been well documented. It may come as an impurity in ethanol^36^ or contamination in supply chains, possibly indicating insufficient purification during emergency production.

From a broader perspective, the USA and Canada demonstrated the highest frequency and variety of contaminants. In contrast, Singapore exhibited fewer contaminants comparatively, indicating stricter product regulations. Nepal, on the other hand, showed the lowest number of contaminants, limited to methanol and inconsistency in ethanol. However, the absence of other compounds could be due to limited sampling or underreporting.

Subpotent products were especially concerning in Nepal (48.7% of records) and Canada (26.7%). A sanitizer with less than the minimum ethanol or isopropanol concentration may not inactivate pathogens, so users receive a false sense of protection. In Nepal, where laboratory capacity is limited, regulators may have detected mainly the failures that are easiest to measure, such as alcohol content. A larger share of records in Nepal and Singapore may also reflect different surveillance approaches rather than different contamination levels.

The USA and Canada had the highest number and widest range of reasons, which is probably related to the size of their markets, to their more active and detailed public reporting, and to the rapid regulatory flexibility they introduced during the emergency. Singapore and Nepal published fewer records, and comparisons between countries should not be over-interpreted. Raw counts do not account for market size, population, or the number of registered products. Because many products are traded internationally, and because the Nepalese market contains many imported products, these national records are likely to reflect global supply-chain problems as well as domestic ones.

This study highlights public health hazards posed by inadequate oversight in healthcare-related products like hand sanitizers, especially in emergency-catalyzed situations like the COVID-19 pandemic. Hazardous contaminants like methanol, acetaldehyde, and subpotency in ethanol, which were frequently observed, are a serious threat to consumers. Failure of manufacturers to maintain compliance with the authorities reveals issues of accountability and a need for regulatory preparedness. To deal with such crisis-driven emergencies, there must be an emergency regulatory pathway that maintains robust and fast analytical verification. Distributed yet certified manufacturing with stern quality monitoring is strongly recommended to effectively deal with such challenges. Any relaxation of manufacturing standards should be accompanied by enforceable post-market obligations, such as a legal duty to respond to regulators, and by a way to identify and delist non-responders quickly. Public messages should stress that sanitizer must never be swallowed, should be stored out of the reach of children, and should be checked for registration and labelling. Finally, distributed manufacturing under certified quality systems has been proposed as a way of expanding supply safely in an emergency; our data are consistent with this idea but do not test it.

## Conclusion

The recall data across Nepal, Canada, Singapore, and the USA underscore a global vulnerability in ensuring the safety and quality of ABHSs during peak demand, as intensified due to the COVID-19 pandemic. The presence of toxic chemical contaminants such as methanol, acetaldehyde, and benzene, along with formulation inconsistencies, indicates manufacturing faults, while issues like improper labelling and circulation of unregistered products reflect regulatory shortcomings. These contaminants pose significant toxicological risks, ranging from acute poisoning to chronic health conditions. While Nepal and Singapore show targeted recall patterns, basically due to the presence of toxic contaminants, Canada and the USA demonstrated a broader regulatory approach reaching up to mislabeling and failure to responding to the authorities. These findings highlight the importance of strong regulatory infrastructures and robust enforcement, especially during public health emergencies. Hand sanitizer recall records from the USA, Canada, Singapore, and Nepal show that the sudden growth in demand during COVID-19 was followed by substantial quality and compliance failures. The regulatory agencies published different types of records, with different criteria and levels of detail, so counts are not strictly comparable. The recall records measure regulatory action, not the prevalence of contamination or of harm. The products that were never tested are not represented. Each record was assigned to a single reason category, which may underestimate the frequency of secondary reasons such as methanol contamination in products also cited for subpotency. We did not analyze contaminant concentrations, manufacturers or product origin as the information was not available, which would give a more detailed picture of risk.

## Supporting information

Rawdata _Canada

Rawdata Nepal

Rawdata _Singapore

Rawdata _USA

## Data Availability

All data produced in the present work are contained in the manuscript.

## Declarations

### Ethical approval

Not Applicable

### Data availability

Data are provided in the main text and in supplementary information.

### Author contribution

Ramdeep Shrestha: Methodology, data curation, formal analysis, writing – original draft

Kriti Khanal: Methodology, formal analysis, writing – review and revision

Astha Neupane: Methodology, formal analysis, writing – review and revision

Basant Giri: Conceptualization, supervision, visualization, writing – review and revision

## References

1. Centers for Disease Control and Prevention (CDC). Hand Hygiene Recommendations. Emerging Infectious Diseases. 2020;26(7):1592–1595.

2. Tarigopula S, Thadipatri R, Mahammed N. A Review on Hand Sanitizers: Types of Hand Sanitizers, Mechanism of action, Toxicity and Adverse Effects COVID-19. Cross Current International Journal of Economics, Management and Media Studies. 2021;3(7):85–91. doi:10.36344/ccijemms.2021.v03i07.002

3. Prajapati P, Desai H, Chandarana C. Hand sanitizers as a preventive measure in COVID-19 pandemic, its characteristics, and harmful effects: a review. Journal of the Egyptian Public Health Association. 2022;97(1). doi:10.1186/s42506-021-00094-x

4. Kratzel A, Todt D, V’kovski P, et al. Inactivation of severe acute respiratory syndrome coronavirus 2 by WHO-recommended hand rub formulations and alcohols. Emerging infectious diseases. 2020;26(7):1592.

5. Golin AP, Choi D, Ghahary A. Hand sanitizers: A review of ingredients, mechanisms of action, modes of delivery, and efficacy against coronaviruses. American Journal of Infection Control. 2020;48(9):1062–1067. doi:10.1016/j.ajic.2020.06.182

6. Mcdonnell G, Russell AD. Antiseptics and Disinfectants: Activity, Action, and Resistance. Clinical Microbiology Reviews. 1999;12(1):147. doi:10.1128/CMR.12.1.147

7. Aodah AH, Bakr AA, Booq RY, et al. Preparation and evaluation of benzalkonium chloride hand sanitizer as a potential alternative for alcohol-based hand gels. Saudi Pharmaceutical Journal. 2021;29(8):807–814. doi:10.1016/J.JSPS.2021.06.002

8. Cucinotta D, Vanelli M. WHO Declares COVID-19 a Pandemic. Acta Biomedica Atenei Parmensis. 2020;91(1):157–160. doi:10.23750/abm.v91i1.9397

9. What Ever Happened to Hand-Sanitizer Startups From the Pandemic? - WSJ. Accessed May 19, 2025. https://www.wsj.com/business/entrepreneurship/hand-sanitizer-companies-2b153270?utm_source=chatgpt.com

10. Hand Sanitizer Market Size, Trends & Statistics Report, 2032. Accessed May 19, 2025. https://www.fortunebusinessinsights.com/hand-sanitizer-market-102710

11. Hand Sanitizer - Nepal | Statista Market Forecast. Accessed May 19, 2025. https://www.statista.com/outlook/hmo/otc-pharmaceuticals/hand-sanitizer/nepal?utm_source=chatgpt.com

12. Methanol and 1-Propanol Contamination in Certain Hand Sanitizer Products | Pharmacy Services | University of Utah Health. Accessed March 26, 2025. https://pharmacyservices.utah.edu/alerts/2020/08/methanol-and-1-propanol-contamination-certain-hand-sanitizer-products

13. Valisure. Accessed March 26, 2025. https://www.valisure.com/valisure-newsroom/valisure-detects-benzene-in-hand-sanitizers

14. Methanol: toxicological overview - GOV.UK. Accessed March 26, 2025. https://www.gov.uk/government/publications/methanol-properties-incident-management-and-toxicology/methanol-toxicological-overview

15. Ashurst J V., Schaffer DH, Nappe TM. Methanol Toxicity. StatPearls. Published online February 6, 2025.

16. Overbeek DL, Watson CJ, Castañeda NR, Ganetsky M. A Geographically Distinct Case of Fatal Methanol Toxicity from Ingestion of a Contaminated Hand Sanitizer Product During the COVID-19 Pandemic. J Med Toxicol. 2021;17(2):218–221. doi:10.1007/s13181-021-00824-9

17. FDA In Brief: FDA Withdrawing Temporary Guidances for Alcohol-Based Hand Sanitizers | FDA. Accessed April 3, 2025. https://www.fda.gov/news-events/press-announcements/fda-brief-fda-withdrawing-temporary-guidances-alcohol-based-hand-sanitizers

18. 2021 Compliance Monitoring Project: Review of ethanol-based hand sanitizers - Canada.ca. Accessed April 3, 2025. https://www.canada.ca/en/health-canada/services/inspecting-monitoring-drug-health-products/compliance-monitoring-reports/2021-2022-review-ethanol-based-hand-sanitizers.html

19. Technical-grade ethanol for the manufacture of hand sanitizers and hard-surface disinfectants during the COVID-19 pandemic: Risk assessment summary report - Canada.ca. Accessed April 4, 2025. https://www.canada.ca/en/health-canada/services/drugs-health-products/natural-non-prescription/legislation-guidelines/covid19-technical-grade-ethanol-hand-sanitizer/risk-assessmentsummary-report.html

20. Staake MD, Goodson FS, Howe GW, et al. Quantitation of Methanol, Ethanol and Isopropanol in Gel Hand Sanitizer Products by GC-FID. 2020;62:92612.

21. Product Recalls, Including Removals and Corrections: Guidance for Industry.

22. Neupane A, Bastakoti M, Tamang S, Giri B. Review of drug recalls and quality of pharmaceutical products in Nepal. BMJ open. 2022;12(7):e053479.

23. Singapore Hand Sanitizer Market | Trends, Share & Size 2031. Accessed May 20, 2025. https://www.6wresearch.com/industry-report/singapore-hand-sanitizer-market-2020-2026?utm_source=chatgpt.com

24. FDA updates on hand sanitizers consumers should not use | FDA. Accessed June 9, 2025. https://www.fda.gov/drugs/drug-safety-and-availability/fda-updates-hand-sanitizers-consumers-should-not-use

25. Coronavirus (COVID-19) Update: FDA Takes Action to Place All Alcohol-Based Hand Sanitizers from Mexico on Import Alert to Help Prevent Entry of Violative and Potentially Dangerous Products into U.S., Protect U.S. Consumers | FDA. Accessed May 20, 2025. https://www.fda.gov/news-events/press-announcements/coronavirus-covid-19-update-fda-takes-action-place-all-alcohol-based-hand-sanitizers-mexico-import

26. Updated: More than 200 hand sanitizers listed on FDA’s ‘do-not-use’ list. Accessed May 20, 2025. https://www.clickondetroit.com/consumer/help-me-hank/2020/10/14/updated-more-than-200-hand-sanitizers-listed-on-fdas-do-not-use-list/

27. Recall of certain hand sanitizers that may pose health risks (Part 1 - June 17, 2020 to March 24, 2021) - Canada.ca. Accessed May 20, 2025. https://recalls-rappels.canada.ca/en/alert-recall/recall-certain-hand-sanitizers-may-pose-health-risks-part-1-june-17-2020-march-24-2021

28. Health Canada warns about certain hand sanitizers that may pose health risks (Part 2 – March 31, 2021 to present) - Canada.ca. Accessed May 20, 2025. https://recalls-rappels.canada.ca/en/alert-recall/health-canada-warns-about-certain-hand-sanitizers-may-pose-health-risks-part-2-march

29. Arasteh P, Pakfetrat M, Roozbeh J. A Surge in Methanol Poisoning Amid COVID-19 Pandemic: Why Is This Occurring? The American Journal of the Medical Sciences. 2020;360(2):201. doi:10.1016/J.AMJMS.2020.05.019

30. Nicol AM. Recalls highlight importance of hand sanitizer safety. Environmental Health Review. 2021;64(1):6–10. doi:10.5864/d2021-005

31. Cucinotta D, Vanelli M. WHO declares COVID-19 a pandemic. Acta bio medica: Atenei parmensis. 2020;91(1):157.

32. Yip L, Bixler D, Brooks DE, et al. Serious adverse health events, including death, associated with ingesting alcohol-based hand sanitizers containing methanol—Arizona and New Mexico, May–June 2020. MMWR Morbidity and Mortality Weekly Report. 2020;69. Accessed October 2, 2026. https://www.cdc.gov/mmwr/volumes/69/wr/mm6932e1.htm?utm_source=mp-fotoscapes

33. Tse TJ, Nelson FB, Reaney MJ. Analyses of commercially available alcohol-based hand rubs formulated with compliant and non-compliant ethanol. International journal of environmental research and public health. 2021;18(7):3766.

34. EPA. Acetaldehyde Hazard Summary. Retrieved from https://www.epa.gov/sites/production/files/2016-09/documents/acetaldehyde.pdf. 2000;January(12):0-3.

35. 15th Report on Carcinogens. doi:10.22427/NTP-OTHER-1003

36. Pal VK, Lee S, Naidu M, Lee C, Kannan K. Occurrence of and dermal exposure to benzene, toluene and styrene found in hand sanitizers from the United States. Environment international. 2022;167:107449. doi:10.1016/J.ENVINT.2022.107449

